# A Scoping Review of Approaches for the Detection and Management of Familial Hypercholesterolaemia in Primary Care

**DOI:** 10.1101/2023.07.13.23292548

**Authors:** Abdullah Zafar Khan, Geoff McCombe, Sarah McErlean, Mark Ledwidge, Tom Brett, Walter Cullen, Joe Gallagher

## Abstract

**Background:** Familial Hypercholesterolaemia (FH) is a genetic condition characterised by a lifelong elevation of low-density lipoprotein cholesterol (LDL-c). FH is one of the most common genetic diseases, with an estimated global prevalence of 1 in 250 individuals. However, it is both underdiagnosed and undertreated. Primary care can be a valuable asset for the opportunistic detection and management of FH.

**Aim:** To examine the employed strategies for improving the detection and management of FH in a primary care setting.

**Method:** Six electronic databases (PubMed, The Cochrane Central Register of Controlled Trials, Web of Science, CINAHL, ProQuest, and Scopus) were searched from May – June 2022 for papers published in English following Arksey and O’Malley’s six-stage scoping review process.

**Results:** The initial search identified 1401 articles and a total of 30 studies were included in this review. A diverse range of methods have been studied for improving identification of FH. Three studies examined reduction in patient LDL-c levels from management in primary care. Two thirds of the studies with primary care management had a significant reduction in patient LDL-c levels.

**Conclusion:** The lack of consistency across the diagnostic criteria and the low number of studies addressing the reduction of patient LDL-c levels are major features of this review. Further research should be conducted to evaluate the effectiveness of the approaches for improving the detection and management of adult patients with FH in a primary care setting.

## Introduction

Familial hypercholesterolaemia (FH) is an autosomal dominant condition characterised by a severe and lifelong elevation of low-density lipoprotein cholesterol (LDL-c). FH has a prevalence of 1 in 250 in the general population, making FH the most common genetic lipid disorder ^1, 2^. Patients with elevated levels of LDL-cholesterol have a significantly higher risk of atherosclerotic cardiovascular disease (ASCVD) due to the cumulative exposure to elevated LDL-cholesterol from birth, compared to those without FH^1–4^.

The World Health Organisation (WHO) has considered FH a “public health priority” since 1998, and advocated for improved screening, early diagnosis and timely initiation of lipid-lowering medications ^1^. FH also meets the WHO criteria for systematic screening and it is recognised as a tier 1 genetic disorder by the U.S Centre for Disease Control (CDC) ^5, 6^. Although some countries have included genetic screening programs in children ^7^ there is still a major problem in the detection and management of FH in adults.

FH remains underdiagnosed and undertreated with approximately 90% of patients remaining undiagnosed globally ^1, 8^. Furthermore, a significant proportion of treated patients do not attain guideline recommended LDL-cholesterol targets ^9–11^. Early diagnosis with lifestyle changes and lipid-lowering therapy can have a significant impact on the reduction of total cholesterol and the early development of ASCVD ^12–14^.

Primary care physicians are ideally placed to assist in the detection and management of FH before it has a pathological manifestation ^15–17^. Less pathological or asymptomatic phenotypes of FH are more commonly seen in primary care compared to specialist care ^18^. Because of its ease of access and frequent patient contact, primary care can be a valuable asset for providing opportunistic screening, especially to those patients who have family history of early ASCVD^15^.

FH is a global health priority and the existing gaps in healthcare need to be addressed to reduce the global burden of FH. The major gap in the healthcare of FH is its sub-optimal detection and management. To address this healthcare gap, we aim to examine the available literature on the strategies to improve the detection and management of FH in adult patients in a primary care setting.

## Methods

A scoping review methodology was chosen to gain a comprehensive overview of the literature in relation to strategies which aim to improve the detection and management of FH in a primary care setting. The scoping review was conducted from May to June 2022, using the six-stage framework described by Arksey and O’Malley ^19^ to collate existing literature, identify key findings and outline current research gaps in this area.

### Stage 1: Identifying the research question

FH is recognised as being common and treatable, but detecting and managing the condition can be challenging. Due to the significant role of primary care in the detection and management of FH in the community, strategies which could improve patient care have been widely researched. Therefore, the objective of this scoping review is to examine the literature for effective strategies which could be implemented to improve detection and management of FH in primary care. We formulated the following research question: “What strategies have been examined to improve the detection and management of familial hypercholesterolaemia in a primary care setting?”

### Stage 2: Identifying relevant studies

A preliminary search of key databases was performed, using multiple search terms to create a reading list. From this, keywords were identified and medical subject heading (MeSH) terms were generated. The electronic databases used in the searches were “PubMed”, the “Cochrane Central Register of Controlled Trials”, “Web of Science”, “CINAHL”, “ProQuest”, and the “Scopus” database (Includes Embase and MEDLINE). The search terms were grouped, with results requiring reference to one or more search term in the following categories: “Detection/Management”, “Familial Hypercholesterolaemia”, and “Primary Care” (Figure 1). Additional articles of relevance were identified by ‘hand-searching’ references that were found in the databases mentioned above.

**Fig 1.**
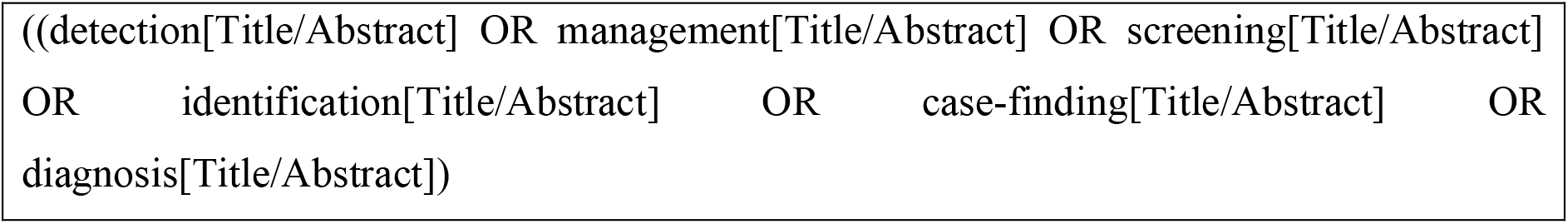
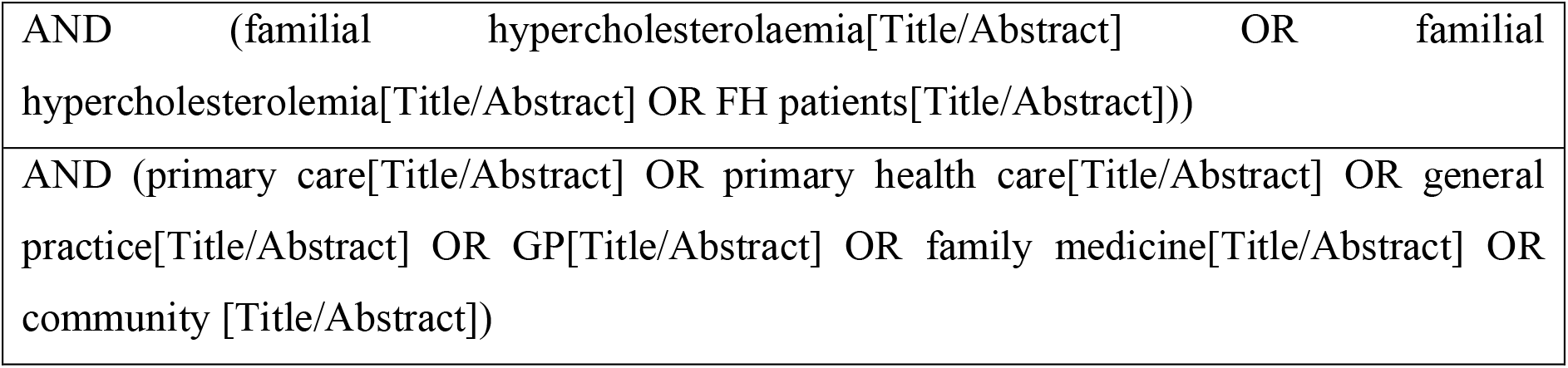
Keywords included in our search strategy, formatted for PubMed

### Stage 3: Study selection

The search identified a total of 1401 citations, of which 39 studies were identified as potentially eligible based on title and abstract screening. Following full-text screening, 30 studies were eligible for inclusion into the review. The ‘Preferred Reporting Items for Systematic Reviews and Meta-Analyses (PRISMA)’ flow diagram below (Figure 2) outlines the selection process.

**Fig 2.**
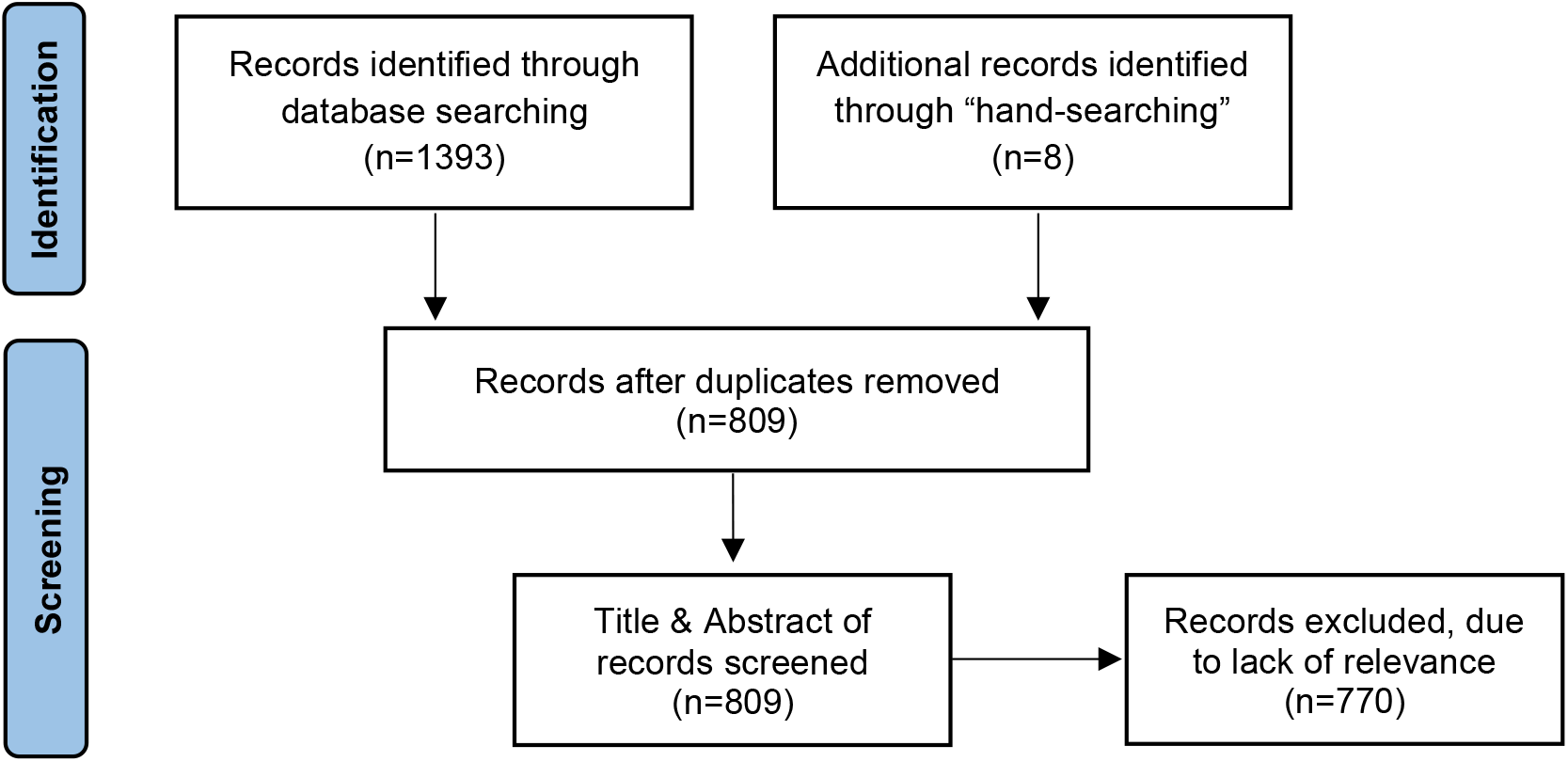
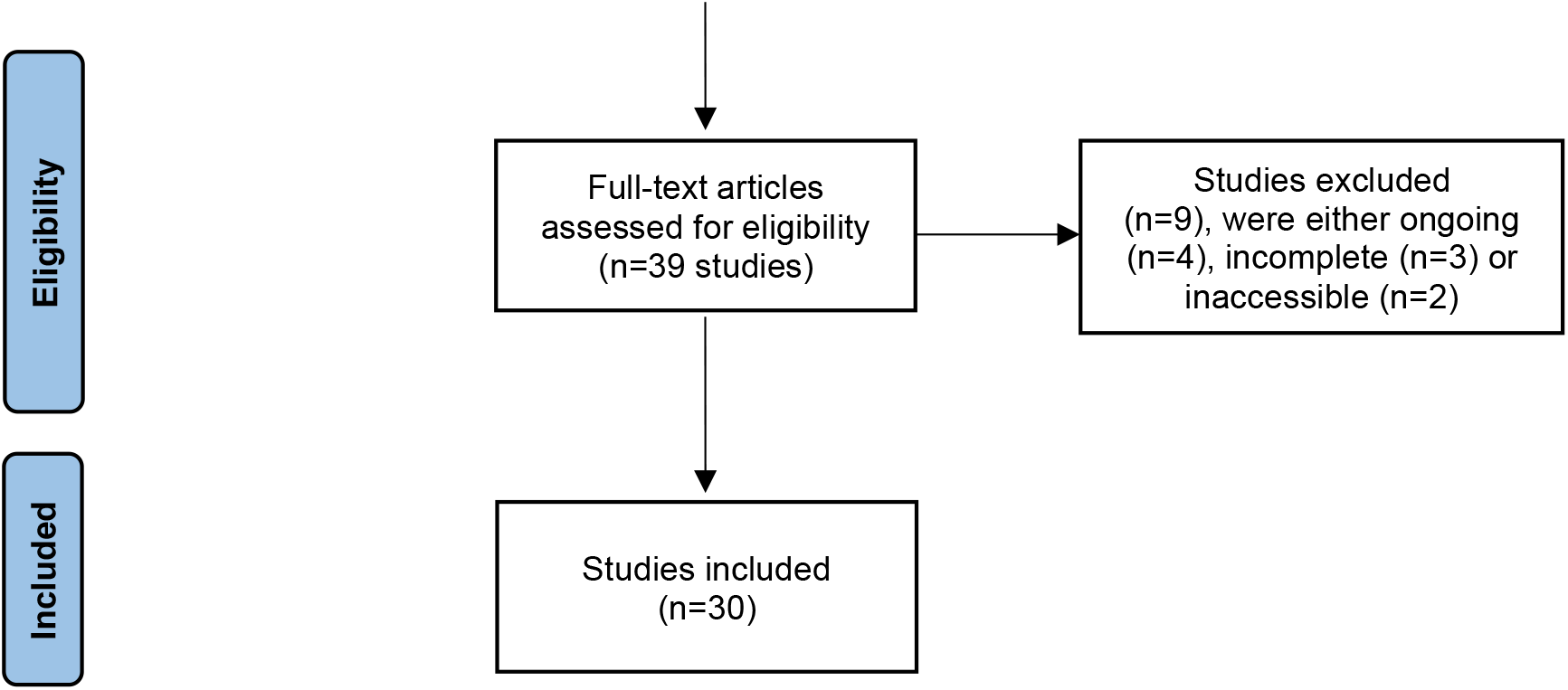
PRISMA Flow diagram

We included any studies that aimed to systematically identify adults with probable or definite FH in a primary care setting. Studies with adult participants of any age from the general population were considered. Studies which included both adults and children were eligible for inclusion if the adult population was separately identified. Community and healthcare system studies that were conducted in a non-specialist setting were also eligible for inclusion. Studies were included if they were published in English and if the full article was available. Studies were excluded due to lack of relevance if they did not aim to systematically identify people with probable or definite FH in a non-specialist setting Findings were reviewed by two other reviewers, and a finalised list of studies was agreed.

### Stage 4: Charting the data

Once all relevant articles were identified (n=30), to facilitate comparison and thematic analysis, the following data were charted from the articles (Table 1):

- First author & year of publication
- Study title
- Study population
- Journal/publication
- Study location
- Study aim/topic
- Strategy employed
- Study design
- Outcome measures
- Major findings

**Table 1.** Studies that aimed to systematically identify adult patients with familial hypercholesterolaemia in a primary care setting

| Primary care-based studies for the improved identification of FH w/o direct involvement of a community laboratory |  |  |  |  |  |  |  |  |  |
| --- | --- | --- | --- | --- | --- | --- | --- | --- | --- |
| Author, year | Study title | Study population | Journal/ publication | Location | Study aim/ topic | Strategy employed | Study design | Outcome measures | Major findings |
| Steyn *et al.* 1998<sup>35</sup> | Detection and measurement of hypercholesterolaemia in South Africans attending general practitioners in private practice—The cholesterol monitor | Patients seen by 200 private general practitioners in South Africa on two survey periods in 1993 and 1994 (n= 12,842) | South African Medical Journal (1998), Vol. 88, Issue 12 | South Africa | Report data on the detection and management of hypercholesterolaemia in patients attending general practitioners in private practice in South Africa | The frequency of cholesterol testing and the level at which active therapeutic intervention occurred at medical practices were monitored over two 5-day monitoring periods in 1993 and 1994 | Cross-sectional survey | Patients identified with possible or definite FH using a medical survey | 12,842 patients were seen by the 200 private practice GPs. Only 3.1% of the patients were reported to have familial hypercholesterolaemia (FH) and 12.8% were reported to have a family history of CHD. 5.4% of the FH patients had TC levels below 5.0 mmol/L, while only 24% of those patients who suffered from CHD had similarly controlled TC levels. Almost 20% of the FH patients had TC levels above 8.0 mmol/l |
| Gray *et al.* 2008<sup>36</sup> | Identifying patients with familial hypercholesterolaemia in primary care: An informatics-based approach in one primary care centre | Patients from a single primary care centre in South London (n= 12,100) | Heart (2008), Vol. 94, Issue 6 | London, UK | Assess the utility of combined computer- and notes-based searches in identifying index cases of FH in primary care, and to uncover the degree of case overlap with secondary care | Four computer-based search strategies were chosen. Selected information such as, highest-recorded cholesterol and physical stigmata, were reviewed by a general practitioner and consultant lipidologist to give a DLCNC score for the probability of FH | Non-randomised, non-controlled, intervention study | Patients identified with possible or definite FH using the DLCNC, Referral to specialist | 402/12,100 (3.3%) patients had a Dutch score high enough to require a notes review. 12 cases of definite FH were found, of whom two were unknown to the practice. Eight probable cases were found, seven of whom were previously unknown. 216/402 (54%) patients scored as possible cases. After specialist review 47/216 (21.8%) patients would merit recalling for a detailed family history and xanthoma examination |
| Kirke *et al.* 2015<sup>37</sup> | Systematic Detection of Familial Hypercholesterolemia in Primary Health Care: A Community Based Prospective Study of Three Methods | Patient records from multiple community databases between January 2010 and December 2012 (n= 94,379) | Heart, Lung and Circulation (2015), Vol. 24, Issue 3 | Western Australia, Australia | Report a prospective study of three methods of case detection using pre-existing primary health care services in one community | Phase 1: Initial screening in primary healthcare settings. Phase 2: Case detection in a primary care setting by a research nurse and general practitioner. Phase 3: Specialist follow-up of high-risk cases | Non-randomised, non-controlled, pre- and post-intervention study | Patients identified with possible or definite FH using the DLCNC and genetic testing, Referral to specialist | 1316 participants underwent detailed assessment for FH. The proportion of at risk people identified for further assessment was in decreasing order: GP (659/2494, 26.4%), workplace assessment (60/268, 22.4%) and pathology database (597/4517, 13.2%) p<0.001. Eighty-six (6.5%) were identified as clinical FH (DLCNC>5) of which 59 had genetic testing and 11/59, 18.6%, were confirmed to have a mutation causing FH. Pathology database detected the greatest number of clinical FH (51/86, 59.3%) and mutation positive participants (8/11, 72.7%) |
| Green *et al.* 2016<sup>27</sup> | Improving detection of familial hypercholesterolemia in primary care using electronic audit and nurse-led clinics | Patients from 56 general practices in the UK (n= 290,000) | Journal of Evaluation of Clinical Practice (2016) Vol. 22, Issue 3 | UK | Assess whether a clinical decision support software in combination with nurse-led clinic can improve the detection of FH in primary care | The first stage was a systematic audit of electronic medical records within GP practices, identifying all patients diagnosed with FH or possible FH and adult patients with a recorded total cholesterol of >7.5 mmol/L or LDL-C ≥5.0 mmol/L. The second stage included a nurse-led clinic to screen more intensely for FH index cases | Non-randomised, non-controlled, pre- and post-intervention study | Possible or definite diagnosis of FH using the Simon-Broome criteria | The baseline prevalence of FH within the study population was 0.13% (1 in 750 persons). After 2 years, the recorded prevalence of diagnosed FH increased by 0.09% to 0.22% (1 in 450 persons). The nurse-led clinic ran for 9 months (October 2013-July 2014) and during this time, the recorded prevalence of patients diagnosed with FH increased to 0.28% (1 in 357 persons) and the prevalence of patients 'at risk and unscreened' reduced from 0.58% to 0.14% |
| Vickery *et al.* 2017<sup>38</sup> | Increasing the Detection of Familial Hypercholesterolemia Using General Practice Electronic Databases | Active patients (≥1 consultation in last two years) from five general practices in Perth, Australia (n= 157,290) | Heart, Lung and Circulation (2017), Vol, 26, Issue 5 | Perth, Australia | Determine whether a simple electronic extraction tool can increase detection of FH in general practice | An extraction tool applied to general practice electronic health records (EHR) to screen for FH, total cholesterol and low density lipoprotein cholesterol (LDL-c) levels in association with entered diagnostic criteria and demographic data | Cross-sectional study | Patients identified with possible or definite FH using LDL-c thresholds | Of 157,290 active patients examined, 0.7% (n=1081) had an LDL-c >5.0 mmol/L representing 1 in 146 of active patients. An additional 0.8% (n=1276) patients were at possible risk of FH. Of those with an LDL-c>5.0 mmol/L, 43.7% had no record of being prescribed statins |
| Casula *et al.* 2017<sup>29</sup> | Detection of familial hypercholesterolemia in patients from a general practice database | Data was collected by more than 600 Italian GPs across 10 Italian regions of patients with measurement of LDL-c (n= 162,864) | Atherosclerosis Supplements (2017), Vol. 29 | Italy | Present a method to improve detection and to enhance awareness of FH in primary care using GP electronic health records | A partial assessment of the DLCNC score using the data that was available. Also determined the prevalence of possible FH based on age-specific LDL-cholesterol thresholds employed by the diagnostic criteria of MED-PED and the non-age adjusted cut-off point (LDL-C ≥190 mg/dL) adopted by the Italian Medicines Agency (AIFA) | Cross-sectional study | Identification of patients with possible or definite FH using the DLCNC, the MED-PED criteria and LDL-c thresholds | Data on LDL-c was available for 162,864 subjects. Mean LDL-C levels were 124.3 mg/dL for non-treated subjects and 106.4 mg/dL for statin-treated subjects. The cut-off of LDL-c ≥190 mg/dL yielded a prevalence of 1:34 among non-treated subjects and of 1:29 among statin-treated patients. Using the cut-off of ≥250 mg/dL, the prevalence was 1:1038 among non-treated subjects and 1:369 among statin-treated patients (average = 1:734). According to the stratification proposed by the MED-PED criteria for the general population, the age-specific LDL-cholesterol thresholds a prevalence of 1:1380 among non-treated subjects and 1:540 among statin-treated patients |
| Aref-Eshghi *et al.* 2017<sup>39</sup> | Identification of Dyslipidemic Patients Attending Primary Care Clinics Using Electronic Medical Record | EMRs of patients ≥20 who had a complete lipid profile taken attending primary care clinics in a Canadian city | Journal of Medical Systems (2017), Vol. 41, Issue 3 | St Johns, Canada | Define the optimal algorithm to identify patients with dyslipidaemia using electronic medical records (EMRs) | Application and comparison of six algorithms. Linear discriminate analysis, and bootstrapping were also performed | Non-randomised, non-controlled, intervention study | Patients identified with possible or definite dyslipidaemia, Sensitivity, negative predictive value, | 3460 individuals (80.6%) were identified as having dyslipidaemia according to the 'gold standard'. Lipid levels showed the best results (sensitivity: 84.0%, NPV: 61.0%, Kappa: 0.67, AUC: 0.67). Among all algorithms, the combination of lipid levels with |

|  | (EMR) Data from the Canadian Primary Care Sentinel Surveillance Network (CPCSSN) Database | during 2009–2010 (n= 4382) |  |  |  |  |  | Kappa coefficient and the AUC were calculated for each algorithm | drug therapy (sensitivity: 100.0%, NPV: 98.0%, Kappa: 0.98, AUC: 1.00) and the combination of lipid levels with ICD codes (sensitivity: 94.0%, NPV: 79.0%, Kappa: 0.85, AUC: 0.97) reached the best results |
| --- | --- | --- | --- | --- | --- | --- | --- | --- | --- |
| Mülvestedt et al. 2021<sup>40</sup> | Screening for potential familial hypercholesterolaemia in general practice: an observational study on prevalence and management | All patients from six general practice clinics and a hospital's cardiology department (n= 9652) | British Journal of General Practice (2021) Vol. 5, Issue 2 | Copenhagen, Denmark | Provide knowledge of the prevalence and management of FH in Danish general practice | Individuals were considered in the group of the high LDL-c population ($\geq 5.0$ mmol/l) and in the group of individuals without secondary hypercholesterolaemia. These groups of individuals were then screened for FH using the DLCNC | Cross-sectional study | Individuals identified with possible or definite FH using the DLCNC | 2382 individuals had a lipid measurement available, and 236 of those had an LDL-c $\geq 5.0$ mmol/l. In total, 34 individuals were found to have probable or definite FH (DLCN score $\geq 5$). Only three individuals had been diagnosed and treated with lipid-lowering therapy. Of 236 individuals with high LDL-c, only 25 individuals met their treatment target. 21 individuals were found to have probable or definite FH (1:114 individuals) |
| Studies that used the FAMCAT algorithm |  |  |  |  |  |  |  |  |  |
| Author, year | Study title | Study population | Journal/ publication | Location | Study aim/ topic | Strategy employed | Study design | Outcome measures | Major findings |
| Qureshi et al. 2021<sup>34</sup> | Case-finding and genetic testing for familial hypercholesterolaemia in primary care | Electronic health records of patients with cholesterol readings from 14 UK general practices (n= 86,219) | Heart (2021), Vol. 107, Issue 24 | England, UK | Describe the genetic and lipid profile of patients found at increased risk of FH and the outcomes in those with positive genetic test results | The Familial Hypercholesterolaemia Case Ascertainment Tool, FAMCAT1 was applied to the patient electronic health records. A family history questionnaire was offered and a detailed review of their clinical data was conducted. After review, genetic testing was offered to those that were considered at risk | Cross-sectional study | Identification of patients with possible FH using FAMCAT and genetic testing, Referral to a specialist | From 86,219 patients with cholesterol readings, 3375 were identified as having an increased risk of FH. Genetic testing was completed by 283 patients, newly identifying 16 with genetically confirmed FH and 10 with variants of unknown significance. In a further 153 (54%) patients, the test suggested polygenic hypercholesterolaemia |
| Qureshi *et al.* 2021<sup>31</sup> | Comparing the performance of the novel FAMCAT algorithms and established case-finding criteria for familial hypercholesterolemia in primary care | Electronic health records of patients with cholesterol readings from 14 UK general practices (n= 86,219) | Open Heart (2021) Vol, 8, Issue 2 | England, UK | Evaluate the performance of two different algorithms (FAMCAT1 and FAMCAT2) at 95% specificity, to detect genetically confirmed FH in the general population | FAMCAT1 and FAMCAT2 were both used to screen for patients at risk of FH. As well as the DLCNC, the Simon-Broome criteria, and recommended cholesterol thresholds. Genetic testing was used as the reference of standard. Detection rate, sensitivity and specificity were examined for each case-finding criterion | Non-randomised, non-controlled, intervention study | Identification of patients with possible or definite FH, Detection rate, sensitivity and specificity of each method | At 95% specificity, FAMCAT 1 had a Detection Rate of 27.8% (95% CI 12.5% to 50.9%) with sensitivity of 31.2% (95% CI 11.0% to 58.7%); while FAMCAT 2 had a DR of 45.8% (95% CI 27.9% to 64.9%) with sensitivity of 68.8% (95% CI 41.3% to 89.0%). DLCN score $\geq 6$ points yielded a DR of 35.3% (95% CI 17.3% to 58.7%) and sensitivity of 37.5% (95% CI 15.2% to 64.6%). Using recommended cholesterol thresholds resulted in DR of 28.0% (95% CI 14.3% to 47.6%) with sensitivity of 43.8% (95% CI 19.8% to 70.1%). Simon-Broome criteria had a lower DR of 11.3% (95% CI 6.0% to 20.0%) and specificity 70.9% (95% CI 64.8% to 76.5%) but higher sensitivity of 56.3% (95% CI 29.9% to 80.2%) |
| Ingoe *et al.* 2021<sup>28</sup> | Improving the identification of patients with a genetic diagnosis of familial hypercholesterolemia in primary care: A strategy to achieve the NHS long term plan | Patients from nine UK general practices were included for screening (n = 94,444) | Atherosclerosis (2021) Vol. 325 | UK | Validate a nurse-led process using electronic health records to identify those at risk of familial hypercholesterolemia (FH) for genetic diagnosis in primary care | Phase 1 of screening used the FAMCAT algorithm. Phase 2 of screening used a modified algorithm which was based on the NICE CG71 guidelines and the DLCN criteria. Patients identified as "very high risk" by both algorithms were triaged by FH specialist nurses for genetic testing | Non-randomised, non-controlled, intervention study | Patients identified as "very high risk" or definite FH using two phases of screening and genetic testing, Referral to specialist | 572 patients (0.61%) were identified as being high risk for FH by both algorithms. 63 patients (53%) underwent genetic testing for FH. And 27 of these patients (43%) were positive for FH |

| Carvalho *et al.* 2021<sup>30</sup> | Application of a risk stratification tool for familial hypercholesterolemia in primary care: an observational cross-sectional study in an unselected urban population | All primary care patients aged 18–65 years registered with general practitioners within three Clinical Commissioning Groups in East London (n=777,128) | Heart (2021), Vol. 107 | London, UK | Test application of the FAMCAT algorithm to describe risks of familial hypercholesterolemia (FH) in a large unselected and ethnically diverse primary care cohort | Retrospectively applied the FAMCAT algorithm to routine primary care data and estimated the numbers of possible cases of FH and the potential service implications of subsequent investigation and management | Cross-sectional study | Identifies patients that have a risk of possible or definite diagnosis of FH using FAMCAT | Of the 777,128 patients studied, the FAMCAT score estimated between 11 736 and 23 798 (1.5%-3.1%) individuals were likely to have FH, depending on an assumed FH prevalence of 1 in 250 or 1 in 500, respectively. There was over-representation of individuals of South Asian ethnicity among those likely to have FH, with this cohort making up 41.9%-45.1% of the total estimated cases, a proportion which significantly exceeded their 26% representation in the study population |
| --- | --- | --- | --- | --- | --- | --- | --- | --- | --- |
| Educational sessions with implementation of a computer-based reminder system |  |  |  |  |  |  |  |  |  |
| Author, year | Study title | Study population | Journal/publication | Location | Study aim/ topic | Strategy employed | Study design | Outcome measures | Major findings |
| Qureshi *et al.* 2016<sup>21</sup> | Feasibility of improving identification of familial hypercholesterolemia in general practice: Intervention development study | Patients from six UK general practices (n=45,033) | BMJ Open (2016), Vol. 6, Issue 5 | Nottinghamshire, UK | Assess the feasibility of improving identification of familial hypercholesterolemia (FH) in primary care, and of collecting outcome measures to inform a future trial | There was a one-hour educational session at each recruited practice. Use of opportunistic computer reminders in consultations and postal invitation over 6 months to eligible patients invited to complete a family history questionnaire. Those fulfilling the Simon-Broome criteria for possible FH were invited for GP assessment and referred for specialist definitive diagnosis | Non-randomised, non-controlled, intervention study | Identification of patients with possible FH using the Simon-Broome criteria and genetic testing, Recruitment rate, Referral to specialist care | From 831 eligible patients, 127 (15.3%) were recruited and completed family history questionnaires: 86 (10.7%) through postal invitation and 41 (4.9%) opportunistically. Among the 127 patients, 32 (25.6%) had a possible diagnosis of FH in primary care. Within 6 months of completing recruitment, 7 patients had had specialist assessment confirming 2 patients with definite FH (28.6%), and 5 patients with possible FH (71.4%) |

| Weng *et al.* 2018<sup>22</sup> | Improving identification and management of familial hypercholesterolaemia in primary care: Pre- and post-intervention study | Patients from six UK general practices (n = 45,033) | Atherosclerosis (2018) Vol. 274 | Nottingham, UK | Assess whether a proactive approach to identify possible FH in primary care improves best practice in accord with UK national guideline recommendations on identifying and managing FH | The intervention was a one-hour educational session with general practitioners and practice nurses on case-identification and assessment of FH using Simon-Broome diagnostic criteria recommended in the current NICE guidelines. There was also use of opportunistic computer reminders in consultations | Non-randomised, non-controlled, pre- and post-intervention study | Identification of possible FH using the Simon-Broome criteria, Cholesterol levels, Statins prescribed, Repeat cholesterol tests | The intervention improved best clinical practice in 118 patients consenting to assessment (of 831 eligible patients): repeat cholesterol test (+75.4%, 95% CI 66.9-82.3); family history of heart disease assessed (+35.6%, 95% CI 27.0-44.2); diagnosis of secondary causes (+7.7%, 95% CI 4.1-13.9), examining clinical features (+6.0%, 95% CI 2.9-11.7). For 32 patients diagnosed with possible FH using Simon-Broome criteria, statin prescription significantly improved (18.8%, 95% CI 8.9-35.3), with non-significant mean reductions in cholesterol post-intervention in both total (-2%) and LDL-c (-3%) |
| --- | --- | --- | --- | --- | --- | --- | --- | --- | --- |
| Studies that used the TAR-B-Ex screening tool |  |  |  |  |  |  |  |  |  |
| Author, year | Study title | Study population | Journal/publication | Location | Study aim/ topic | Strategy employed | Study design | Outcome measures | Major findings |
| Troeung *et al.* 2016<sup>23</sup> | A new electronic screening tool for identifying risk of familial hypercholesterolaemia in general practice | All active patients seen at a large general practice in Perth, Western Australia between 2012 and 2014 (n= 3708) | Heart (2016), Vol. 102, Issue 11 | Perth, Australia | Evaluate the performance of a new electronic screening tool (TARB-Ex) in detecting general practice patients at potential risk of familial hypercholesterolaemia (FH) | Retrospective screening for potential FH risk using TAR-B-Ex. Electronic extracts of medical records for patients identified with potential FH risk (defined as DLCNC score ≥5) through TAR-B-Ex were reviewed by a general practitioner (GP) and lipid specialist. High-risk patients were recalled for clinical assessment | Non-randomised, non-controlled, intervention study | Identification of possible or definite FH using the DLCNC and genetic testing; Sensitivity, specificity, positive predictive value, and the negative predictive value were all examined, Screening time, Referral to specialist | 32 patients with DLCNC score ≥5 were identified through electronic screening compared with 22 through GP manual review. Sensitivity was 95.5% (95% CI 77.2% to 99.9%), specificity was 96.7% (95% CI 94.3% to 98.3%), negative predictive accuracy was 99.7% (95% CI 98.3% to 100%) and positive predictive accuracy was 65.6% (95% CI 46.9% to 8%). Electronic screening was completed in 10 min compared with 60 h for GP manual review. 10/32 patients (31%) were considered high risk and recalled for clinical assessment. Six of seven patients (86%) who attended clinical assessment were diagnosed with phenotypic FH on examination |

| Brett *et al.* 2021<sup>24</sup> | Improving detection and management of familial hypercholesterolemia in Australian general practice | Patients that attended 15 general practices ($\geq 1$) in the previous 2 years (n = 232,139) | Heart (2021), Vol. 107 Issue 15 | Across 5 Australian states: Western Australia, New South Wales, Queensland, Victoria, Tasmania | Examine the feasibility of using an electronic screening tool (TARB-Ex) to identify and better manage primary care patients with FH | First stage was TARB-Ex screening, then came manual record review, clinical assessment and information from follow-up consultation | Non-randomised, non-controlled, pre- and post-intervention study | Individuals identified with possible or definite FH using the DLCNC, Cholesterol levels, Lipid-lowering treatment prescription | 1843 patients were identified by TARB-Ex as at potential risk of FH (DLCNC score $\geq 5$). After GP medical record review, 900 of these patients (49%) were confirmed with DLCNC score $\geq 5$ and classified as high-risk of FH. After follow-up, there were significant reductions in plasma total cholesterol ($-9\%$) and LDL-cholesterol ($-16\%$) levels in patients with GP management compared with baseline (untreated LDL-c) ($p < 0.01$) |
| --- | --- | --- | --- | --- | --- | --- | --- | --- | --- |
| Community and healthcare system-based studies for the improved identification of FH w/o direct involvement of a community laboratory |  |  |  |  |  |  |  |  |  |
| Author, year | Study title | Study population | Journal/publication | Location | Study aim/ topic | Strategy employed | Study design | Outcome measures | Major findings |
| Benn *et al.* 2012<sup>41</sup> | Familial hypercholesterolemia in the Danish general population: Prevalence, coronary artery disease, and cholesterol-lowering medication | An unselected, community-based population from the Copenhagen General Population Study (n= 69,016) | The Journal of Clinical Endocrinology and Metabolism (2012), Vol. 97, Issue 11 | Copenhagen, Denmark | Investigate the prevalence of FH and the associations between FH and coronary artery disease and cholesterol-lowering medication in the Copenhagen General Population Study | The diagnostic criteria used was the Dutch Lipid Clinic Network Criteria. Information on coronary artery disease was collected. Plasma concentrations of cholesterol, high-density lipoprotein (HDL)-cholesterol, triglycerides, and glucose were measured | Cross-sectional study | Patients identified as having possible or definite FH using the DLCNC and genetic testing | The prevalence of FH was 0.73% (1:137). The prevalence of coronary artery disease among FH participants was 33%. Only 48% of subjects with FH admitted to taking cholesterol-lowering medication. The odds ratio for coronary artery disease off cholesterol-lowering medication was 13.2 (10.0-17.4) in definite/probable FH compared with non-FH subjects. The corresponding odds ratio for coronary artery disease in FH subjects on cholesterol-lowering medication was 10.3 (7.8-13.8, 95% CI) |
| Safarova *et al.* 2016<sup>42</sup> | Rapid identification of familial hypercholesterolemia from electronic health | Individual lipid levels were extracted from structured laboratory databases from June 21, 1993, to | Journal of Clinical Lipidology (2016), Vol. 10, Issue 5 | Minnesota, USA | Developed an electronic phenotyping algorithm for rapid identification of FH in electronic health records (EHRs) and | A modified numerical score system of the DLCN criteria. Several other variables that were incorporated in the SEARCH electronic phenotyping algorithm | Cohort study | Identification of possible or definite FH diagnosis using the SEARCH algorithm, Cholesterol levels | The SEARCH algorithm identified 32 definite and 391 probable cases with an overall FH prevalence of 0.32% (1:310). Only 55% of the FH cases had a diagnosis code relevant to FH. Mean LDL-c at the time of FH ascertainment was 237 mg/dL; at follow-up, 70% (298 |
| | records: The SEARCH study | December 31, 2014.<br>(n= 131,000) | | | deployed it in the Screening Employees And Residents in the Community for Hypercholesterolemia (SEARCH) study | | | | of 423) of patients were on lipid-lowering treatment with 80% achieving an LDL-C ≤100 mg/dL. Of treated FH patients with premature CHD, only 22% (48/221) achieved an LDL-C ≤70 mg/dL |
| Zamora *et al.* 2017<sup>43</sup> | Familial hypercholesterolemia in a European Mediterranean population—Prevalence and clinical data from 2.5 million primary care patients | Patient records from a Spanish patient information system aged ≥8 years, alive on December 2014, and with at least 1 low-density lipoprotein cholesterol (LDL-c) measurement between 2006 and 2014 (n= 2,554,644) | Journal of Clinical Lipidology (2017), Vol. 11, Issue 4 | Spain | Estimate the prevalence of the FH phenotype (FH-P) and to describe its clinical characteristics in a Mediterranean population | Heterozygous FH-P and homozygous FH-P was defined as an untreated low-density lipoprotein cholesterol plasma concentration threshold. The presence of cardiovascular diseases and risk factors was defined by coded (ICD-9 and ICD-10) medical records from primary care and hospital discharge databases | Cross-sectional study | Patients identified with possible FH using age-adjusted LDL-c thresholds | The age- and sex-standardized prevalence of heterozygous FH-P and homozygous FH-P were 1/192 individuals and 1/425,774 individuals, respectively. Among patients with FH-P aged $>$18 years, cardiovascular disease prevalence was 3.5 times higher than in general population, and CHD prevalence in those aged 35 to 59 years was 4.5 times higher than in those without FH-P. Lipid-lowering therapy was lacking in 13.5% of patients with FH-P, and only 31.6% of men and 22.7% of women were receiving high or very high-intensity lipid-lowering therapy |
| Elis *et al.* 2020<sup>44</sup> | The characteristics of patients with possible familial hypercholesterolemia—screening a large payer/provider healthcare delivery system | Medical records from an Israeli healthcare fund between 31 December 2006 to 1 January 2018 (n= ~4,500,000) | QJM (2020), Vol. 113, Issue 6 | Israel | Applied standard laboratory criteria across a large electronic medical record database to describe cross-sectional population with possible FH | Individuals that met the MED-PED criteria were considered to have possible FH, excluding those with secondary causes. Demographic and clinical characteristics were described to estimate the burden of FH on healthcare | Cross-sectional study | Patients identified with possible FH using the MED-PED criteria | The study cohort included 12,494 subjects out of around 4.5 million. The estimated prevalence in Israel was found to be 1:285. These patients had a significant family history of cardiovascular disease. Most of the patient's LDL-c was not controlled and only 25% were medically treated |
| Chua *et al.* 2021<sup>45</sup> | Familial Hypercholesterolemia in the | Participants attending a Health Screening | Journal of Atherosclerosis and | Malaysia | Report the first nation-wide investigation on the | Blood samples were collected for lipid profiles and glucose | Cross-sectional study | Identifies patients that have a possible or | Out of 5130 recruited community participants, 55 patients were clinically categorised as potential FH, making the |
|  | Malaysian Community: Prevalence, Under-Detection and Under-Treatment | Programme across Malaysia from 2011 to 2019 (n= 5130) | Thrombosis (2021), Vol. 28, Issue 10 |  | prevalence of FH in the Malaysian population, their detection rate, proportion on lipid-lowering treatment and control within the Malaysian community | analyses. Personal and family medical histories were collected by means of assisted questionnaire. Physical examination for tendon xanthomata and premature corneal arcus were conducted on-site. FH was clinically screened using the DLCNC |  | definite diagnosis of FH using the DLCNC | prevalence FH among the community 1:100. Based on current total population of Malaysia (32 million), the estimated number of FH patients in Malaysia is 320,000, while the detection rates are estimated as 0.5%. Lipid-lowering medications were prescribed to 54.5% and 30.5% of potential and possible FH patients, respectively, but none of them achieved the therapeutic LDL-c target |
| Eid *et al.* 2022<sup>46</sup> | Improving Familial Hypercholesterolemia Diagnosis Using an EMR-based Hybrid Diagnostic Model | Patient electronic medical records (EMR) showing any lipid profile from a clinical encounter in a US healthcare system between January 1, 2009, and April 30, 2020 (n = 289,299) | The Journal of Clinical Endocrinology and Metabolism (2022), Vol. 107 Issue 4 | Kentucky, USA | Test the utility of a hybrid diagnostic model to determine FH prevalence and treatment characteristics in the study population | Dynamic EMR-based clinical decision support tool used in combination with the DLCNC or the AHA criteria for diagnosis of FH | Cross-sectional study | Identifies patients that have a possible or definite diagnosis of FH using the DLCNC and the AHA criteria, Referral to specialist | From 264,264 patient records, between 794 and 1571 patients were identified as having FH based on the hybrid diagnostic model, with a prevalence of 1:160 to 1:300. These patients had a higher prevalence of premature CAD (38-58%) than the general population (1.8%) and higher than those having a high CAD risk but no FH (10%). Although most patients were receiving lipid-lowering therapies (LLTs), only 50% were receiving guideline-recommended high-intensity LLT |
| Fasano *et al.* 2022<sup>47</sup> | Search for familial hypercholesterolemia patients in an Italian community: A real-life retrospective study | Individuals in a laboratory database (n= 221,644) and patients with ASCVD (n= 583) who underwent percutaneous coronary angioplasty (PCTA) | Nutrition, Metabolism & Cardiovascular Diseases (2022), Vol. 32, Issue 3 | Italy | Identify and genetically characterize potential FH patients referred to the Lipid Clinic and monitor attainment of treatment goals in identified patients | Monitored the lipid profiles of subjects with LDL-C ≥ 250 mg/dl identified by laboratory survey, PTCA patients and patients from the Lipid Clinic | Cross-sectional study | Identifies patients that have a possible or definite diagnosis of FH using the DLCNC and genetic testing, Referral to a specialist | The laboratory survey identified 1.46% of subjects with LDL-C ≥ 190 mg/dl (1:68) and 0.08% with LDL-C ≥ 250 mg/dl (1:125). Probable/definite FH was suspected in 3% of PTCA patients. Molecularly-confirmed FH was found in 44% of subjects with clinical suspicion of FH. The 50% LDL-c reduction target was achieved by 70.6% of subjects with clinical suspicion of FH Only 18.5% of PTCA patients reached the LDL-c < 55 mg/dl target |

| Studies that directly involved community laboratories |  |  |  |  |  |  |  |  |  |
| --- | --- | --- | --- | --- | --- | --- | --- | --- | --- |
| Author, year | Study title | Study population | Journal/publication | Location | Study aim/ topic | Strategy employed | Study design | Outcome measures | Major findings |
| Bell *et al.* 2012<sup>48</sup> | Opportunistic screening for familial hypercholesterolaemia via a community laboratory | Individuals with a cholesterol result from a private community laboratory in Western Australia between 1<sup>st</sup> May 2010 to 30<sup>th</sup> April 2011 (n= 84,823) | Annals of Clinical Biochemistry (2012) Vol. 49, Issue 6 | Western Australia, Australia | Determine the ability of a community laboratory to screen for individuals with potential FH | The prevalence of possible FH was based on LDL-cholesterol thresholds employed by the Make Early Diagnosis-Prevent Early Death (MED-PED), the Simon Broome Registry and the Dutch Lipid Clinic Network Criteria | Cross-sectional study | Possible or definite diagnosis of FH using LDL-c thresholds | During this period, 84,823 patients had 99,467 serum LDL-cholesterol measurements, with 91.8% requested by general practitioners. The prevalence of FH based on an LDL-cholesterol $\geq 250$ mg/dL, the 99.75th percentile, was 1:398 in this sample population, only the MED-PED LDL-cholesterol criteria gave a similar prevalence of 1:482 |
| Bell *et al.* 2013<sup>25</sup> | Impact of interpretative commenting on lipid profiles in people at high risk of familial hypercholesterolaemia | Individuals with an LDL-c of $\geq 6.5$ mmol/L on lipid profile requested by a GP between 23<sup>rd</sup> June and 19<sup>th</sup> October 2010 | Clinica Chimica Acta (2013), Vol. 422 | Western Australia, Australia | Investigate whether interpretative commenting on lipid profiles could improve FH detection and treatment | Interpretative comments were added to the lipid results with the assistance of an expert system, with all comments reviewed by a chemical pathologist | Case-historical control study | Identification of patients with possible FH using LDL-c thresholds, Cholesterol levels, Referral to a specialist | Interpretative commenting was associated with a significant additional LDL-c reduction (0.7mmol/L) compared with controls. A minority of individuals (3/26, 11.5%) received a specialist referral |
| Bell *et al.* 2014<sup>16</sup> | Can Patients be Accurately Assessed for Familial Hypercholesterolaemia in Primary Care? | Individuals at risk of FH were identified by either the laboratory highlighting individuals with elevated LDL-c, or by using an informatics tool to search general practice databases (n= 153) | Heart, Lung and Circulation (2014), Vol. 23, Issue 12 | Western Australia, Australia | Investigate whether individuals with FH could be accurately identified in primary care | Individuals at risk of FH were identified by either the laboratory highlighting individuals with elevated LDL-c, or by using an informatics tool to search general practice databases. DLCNC scores assessed by GPs, were compared with the DLCNC assessed by specialists using primary care data in 153 individuals. 30 | Non-randomised, non-controlled, intervention study | Identification of patients with possible or definite FH using the DLCNC and genetic testing, Referral to a specialist | GPs correctly classified 39 (86.7%) individuals with 'clinical FH', and 32 (94%) with 'unlikely FH' relative to specialists. Lin's concordance correlation coefficient was high (0.832 (0.783 - 0.881), $p < 0.001$) between specialist and GPs, with an overall agreement of 83.6%, 0.744 (0.642 - 0.831). After specialist review, 15 individuals (50%) were diagnosed with clinical FH, four (26.7%) had FH mutations. GPs correctly classified 12 (80%) of these individuals with clinical FH |
| | | | | | | individuals with DLCNC score $\geq 4$ underwent specialist review and genetic testing | | | |
| Bell *et al.* 2014<sup>49</sup> | Detecting familial hypercholesterolemia in the community: Impact of a telephone call from a chemical pathologist to the requesting general practitioner | Individuals that were having their LDL-cholesterol measured by a private community laboratory in Western Australia, at the request of a GP. The Intervention group consisted of the first 100 individuals whose GP's answered the call between 1<sup>st</sup> November 2010 and 6<sup>th</sup> October 2011 (n=100) | Atherosclerosis (2014), Vol. 234, Issue 2 | Western Australia, Australia | Determine whether a telephone call from a chemical pathologist to the requesting general practitioner (GP) of individuals at high risk of familial hypercholesterolemia (FH) increases specialist referral and detection of FH | All laboratory reports (cases and controls) received interpretative comments highlighting FH. In addition, the cases' GPs received a telephone call from the chemical pathologist to highlight their patient's risk of FH and suggest specialist referral, the controls' GPs were not telephoned | Case-historical control study | Identification of patients with possible or definite FH using an LDL-c threshold and genetic testing, Referral to a specialist | After 12 months follow-up, 27 (27%) cases were referred to clinic compared with 4 (4%) controls ($p < 0.0001$). 25 cases were reviewed at clinic, 12 (48%) had definite FH and 18 (72%) had probable or definite FH according to the DLCNC, 2 cases did not attend their clinic appointments. Genetic testing was performed in 23 individuals: 7 (30%) had pathogenic FH mutations. Genotypic cascade screening of 4 kindreds from the intervention group detected an additional 7 individuals with FH and excluded 5 mutation-negative family members |
| Bell *et al.* 2015<sup>50</sup> | The potential role of an expert computer system to augment the opportunistic detection of individuals with familial hypercholesterolemia from a community laboratory | Patients who had lipid profiles requested from a private community laboratory in Western Australia, between the 1st of May 2010 and the 30th of April 2011 (n= 84,823) | Clinica Chima Acta (2015), Vol. 448 | Western Australia, Australia | Determine if an expert system (ES) at a community laboratory could identify information relevant for estimating an individual's likelihood of FH using the DLCNC | The ES was used to retrospectively search a database consisting of laboratory results and clinical details. The DLCNC was used to estimate an individual's risk of FH | Non-randomised, non-controlled, intervention study | Identify individuals at risk of FH using the DLCNC | 84,823 individuals had $\geq 1$ LDL-cholesterol request with data available on 84,083 (99.1%). Clinical details were provided on 71,282 (84.8%) individuals. History relevant to the DLCNC was present in 883 (1.1%) individuals, with premature CVD and non-cardiac vascular disease present in 177 and 64 individuals, respectively. Statin therapy was reported in 5118 individuals; 112 individuals with a current LDL-cholesterol of $< 6.5$ mmol/L had a previous LDL-cholesterol of $\geq 6.5$ mmol/L |
| Bender *et al.* 2016<sup>26</sup> | Interpretative comments specifically suggesting specialist referral increase the detection of familial hypercholesterolemia | Individuals referred by a GP who were found to have an LDL-cholesterol $\geq 6.5$ mmol/L measured at a private community laboratory in Western Australia, between 1 December 2012 and 1 December 2013 (n= 231) | Pathology (2016), Vol. 48, Issue 5 | Western Australia, Australia | Determine whether specifically recommending referral to the regional Lipid Disorders Clinic (LDC) increased referral and FH detection rates | Interpretative comments were added to the lipid results with the assistance of an expert system with all comments reviewed by one of two chemical pathologists before being issued. All subjects received an interpretative comment that raised FH as a consideration. The cases received an additional recommendation for referral to a lipid specialist | Prospective case-control study | Identify individuals with possible FH using LDL-c thresholds and genetic testing, Referral to a specialist | There were 231 individuals with an LDL-cholesterol $\geq 6.5$ mmol/L; 96 (42%) controls and 135 (58%) cases, of which 99 were fax-cases. Twenty-four (18%) cases were referred to clinic compared with eight (8%) controls (p = 0.035). After specialist review and genetic testing, four probable and four definite FH individuals were detected amongst controls, compared with seven possible, eight probable and nine definite FH amongst cases. Genetic testing was performed in 31 (94%) individuals, 13 (42%) had a causative mutation identified |
| Fath *et al.* 2021<sup>51</sup> | FH ALERT: efficacy of a novel approach to identify patients with familial hypercholesterolemia | Patients aged 60 and below who were being evaluated for elevated LDL-C or total cholesterol (TC) measurements between March 15, 2018 and June 15, 2018 (n = 60,812) | Scientific Reports (2021), Vol. 11 | Bavaria, Germany | Assess whether alerting physicians for the possibility of FH impacted additional diagnostic activity | Participating physicians received an alert letter (AL) once total cholesterol or LDL-c levels exceeded predefined threshold values. The ALs also included scientific information about FH and the further diagnostic options including genetic testing | Non-randomised, non-controlled, intervention study | Possible diagnosis of FH using an LDL-c threshold and genetic testing | ~5% of all tested samples triggered an FH alert which was triggered by 2846 patients. 93 genetic test were sent and 26 genetic tests (28%) were returned, 5 patients (19%) tested positive for FH |

### Stage 5: Collating, summarising and reporting results

An overview of the literature is detailed in Table 1 below, summarising and charting the results. This has been discussed further in the results section.

### Stage 6: Consultation

In line with recommendations by Levac et al ^20^, studies were also included and excluded according to advice received during consultation with experts in the field of familial hypercholesterolaemia research in primary care

## Results

The initial database searches identified 1401 articles. After 592 duplicates were removed, reviewers screened the remaining 809 articles by title and abstract, during which 770 articles were excluded. 39 studies met the inclusion criteria and were selected for full-text review. Following full-text review, nine studies were excluded due to four studies being ongoing, three studies were incomplete, and two were made inaccessible by the authors. Data was extracted from the final selection consisting of 30 studies which met the eligibility criteria for the review. The search process, as guided by the Preferred Reporting Items for Systematic Reviews and Meta-Analyses (PRISMA), is summarised in Fig. 2. Data was extracted from the final selection consisting of 30 studies which met the eligibility criteria for the review.

### Description of included studies

From the 30 included studies, 13 were cross-sectional studies, nine were non-randomised, non-controlled intervention studies, four were non-randomised, non-controlled, pre- and post-intervention studies, three were case-control studies and one was a cohort study. The majority of studies were conducted in Australia (n=10) and the UK (n=8). The other countries with more than a single study were, Italy (n=2), Denmark (n=2) and the USA (n=2).

### Study populations and settings

Study populations were most commonly individuals that had a measurement of their total cholesterol or LDL-c registered in a clinical or laboratory database (n=15). Other populations included patients from primary care databases (n=10), individuals from a multi-database search (n=2), a community-based population (n=1), individuals in a healthcare fund (n=1), and individuals that attended a health screening programme (n=1). All of these studies took place in a primary care or community setting (n=30).

### Diagnostic criteria

For most of the studies, the Dutch Lipid Clinic Network Criteria (DLCNC) was used to identify possible or definite cases of FH (n= 14). The Simon-Broome criteria (n=4) and the MED-PED criteria (n=2) were also used. LDL-c and total cholesterol thresholds were also successfully implemented in some studies (n=9). Genetic testing was used to confirm the FH diagnosis or to identify the FH-causing mutation (n=12).

### Strategies employed

An intervention that was studied in the UK was a one-hour educational session ^21, 22^, which aimed to educate primary care physicians on the identification of FH using the Simon-Broome criteria (n=2). Both of these studies included computer-based reminder messages that prompted the primary care physician to opportunistically assess eligible patients for FH during consultation. TARB-Ex was an electronic extraction tool that specifically screened for FH, accounting for two Australian studies ^23, 24^.

Emphasis on the direct involvement of community laboratories was given importance in many studies (n=7). Two of these studies were based on FH interpretive comments systems ^25, 26^. Some strategies included specialist nurse (n=2) and specialist lipidologist (n=3) involvement in the review process. Specialist nurses triaged potential FH cases for genetic testing and also independently screened for FH index cases ^27, 28^.

### FH prevalence

FH prevalence was estimated with application of the DLCNC (n=4), the MED-PED (n=1), LDL-c thresholds (n=2) and one study used a combination of DLCNC, the MED-PED criteria and LDL-c thresholds ^29^. In all four of the studies that used the DLCNC, the estimated prevalence of probable or definite FH was between 1:100 and 1:137. The study that used the MED-PED criteria estimated the prevalence of probable or definite to be 1:285. Studies that estimated the prevalence of probable or definite FH using the LDL-c threshold of ≥250 mg/dL estimated a prevalence between 1:398 and 1:734. While the FAMCAT algorithm estimated the prevalence of FH to be between 1:250 and 1:500 in a London population ^30^.

### FH diagnosis

The educational session/computer-reminder study conducted by Weng *et al*. ^22^ and the electronic audit/nurse-led clinic model by Green et al. ^27^, both demonstrated an improvement of FH diagnosis when compared to baseline data of usual care. Prevalence studies, electronic extraction tools/searches, the FAMCAT algorithm, and FH alert/comment systems were the other employed strategies that increased FH diagnosis.

Importantly, the novel FAMCAT2 algorithm had a higher detection rate and a higher sensitivity when compared to the FAMCAT1 algorithm, the DLCNC, the Simon-Broome criteria and recommended cholesterol thresholds ^31^.

### Lipid management

There were three studies that included lipid management in a primary care setting. The interpretive comment study produced a significant reduction (-23%, p<0.005) in the LDL-c levels of patients when compared with controls ^25^. The TARB-Ex study by Brett *et al*. also demonstrated significant mean reductions in both plasma cholesterol (-9%, p<0.01) and LDL-cholesterol (-16%, p<0.01) when compared to baseline ^24^. In the computer-reminder study conducted by Weng *et al.* there were many significant clinical improvements including the examination of clinical features, family history assessment and statin prescription ^22^. However, there were non-significant reductions in both plasma cholesterol (-2%) and LDL-cholesterol (-3%).

## Discussion

### Summary

This review aimed to map the literature concerning the detection and management of FH in primary care. Research to date has predominantly been focused on strategies that aim to increase the identification of individuals with probable or definite FH. However, increased identification of probable or definite FH does not imply an improvement in clinical outcomes.

Bell *et al.* found that GP assessment of FH was comparable to specialist assessment ^16^. However, management of FH in the included studies was more commonly with specialist care rather than primary care alone. Twelve of the studies in this review included individual referral to a specialist service. Although specialist care was more favoured than primary care, two thirds of the studies with primary care management of patient LDL-c levels demonstrated a significant reduction when compared with baseline. A minority of studies in this review included baseline data of usual care (n=6).

The computer-reminder study by Weng *et al.* and the interpretive comment study by Bell *et al.* provided two examples of the positive impact that FH alert/comment systems could have on improving clinical practice and reducing patient LDL-c levels ^22, 25^. These systems could be effective supplements for assisting primary care physicians in the detection and management of FH patients. However, these systems should be studied further.

### Strengths and limitations

This review utilised the Arksey and O’Malley’s scoping review framework, and identified significantly more studies than previous reviews. Although we aimed to be comprehensive in our approach, there is a possibility that not all publications relevant to the subject area were identified by the search strategy. In addition, scoping reviews do not include an assessment of study quality as the focus is on covering the whole range of relevant literature. Furthermore, only articles published in English were considered for inclusion into our review, which could have resulted in the exclusion of relevant literature published in other languages. We did not include paediatric screening studies, including genetic screening, as these are not part of routine primary care but these are increasing as population and public health measures.

### Comparison with existing literature

There have been two previous systematic reviews that focused on this topic. In the first review, the authors assessed 30 studies for eligibility and none were included ^32^. In the second review, the authors assessed 29 studies for eligibility and included three into their review ^33^. Both systematic reviews concluded that there was insufficient evidence to determine the most effective method of systematically identifying FH in non-specialist settings. Our scoping review included significantly more articles and a diverse range of methods for FH diagnosis. We presented key information about the included studies in Table 1.

### Implications for research and practice

There were many studies that aimed to systematically identify individuals with FH. However, few studies had baseline data and there was a lack of studies that validated their methods of identification using genetic confirmation (n=12). There was also a lack of consistency between the diagnostic criteria. As a result, there is currently insufficient evidence to inform the most effective approach for the detection and management of FH in primary care. More research needs to be conducted to evaluate the impact that these models can have on the reduction of clinical events. The role of genetic testing in primary care is also evolving and will likely be significant in the future of FH detection and management.

## Conclusion

FH is a common genetic condition, but timely detection and management has significant potential to reduce morbidity and mortality in families. Primary care is ideally placed to undertake this work given its close relationship with families and its success in general cardiovascular disease prevention. Therefore, it is not surprising that a number of models of FH case-finding have been developed and studied. The lack of consistency across the diagnostic criteria and the low number of studies addressing the reduction of patient LDL-c levels are major features of this review. Genetic testing is becoming more accessible and its integration into primary care will require further evaluation. Conditions such as polygenic hypercholesterolaemia can represent a large proportion of possible FH cases and its genetic implications should be studied further ^34^. Further research should be conducted to evaluate the effectiveness of the approaches for improving the detection and management of adult patients with FH in a primary care setting.

## Data Availability

All data produced are available online

## Acknowledgements

No funding was provided for this research. The authors stated that they had no interests which might be perceived as posing a conflict or bias.

